# Causal estimands and target trials for the effect of lag time to treatment of cancer patients

**DOI:** 10.64898/2026.04.07.26350338

**Authors:** Bronner P. Gonçalves, Eduardo L. Franco

## Abstract

Timeliness of therapy initiation is a fundamental determinant of outcomes for many medical conditions, most importantly, cancer. Yet, existing inefficiencies in healthcare systems mean that delays between diagnosis and treatment frequently adversely affect the clinical outcome for cancer patients. Although estimates of effects of lag time to therapy would be informative to policymakers considering resource allocation to minimize delays in oncology, causal methods are seldom explicitly discussed in epidemiologic analyses of these lag times. Here, we propose causal estimands for such studies, and outline the protocol of a target trial that could be emulated with observational data on lag times. To illustrate the application of this approach, we simulate studies of lag time to treatment under two scenarios: one in which indication bias (Waiting Time Paradox) is present and another in which it is absent. Although our discussion focuses on oncologic outcomes, components of the proposed target trial could be adapted to study delays for other medical conditions. We believe that the clarity with which causal questions are posed under the target trial emulation framework would lead to improved quantification of the effects of lag times in oncology, and hence to better informed policy decisions.

## 1. Introduction

The time interval that elapses between the diagnosis of a clinical condition and the initiation of therapy might affect morbidity and mortality outcomes of patients (1). A medical field in which this has been studied in detail is oncology; several studies (2-7) have described different aspects of delays between diagnosis and treatment in cancer patients. This research literature revealed considerable variation in the associations between lag time and oncologic outcomes, and in some settings, shorter time intervals between diagnosis and treatment were associated with higher (rather than, as one would expect, lower) mortality. Although this might be partially related to differences between populations, epidemiologic biases could contribute to these variable results. Specifically, indication bias, whereby patients with more severe disease are prioritized for care, might explain observations of shorter lag times being associated with poorer outcomes; this phenomenon is known as the Waiting Time Paradox, and also occurs for lag times between other steps of cancer care (8-10), for example, for the lag time from symptom onset to diagnosis.

The decision of whether to allocate or not more resources to minimize lag times between steps of oncologic care should be informed by studies that ask precise causal questions. And yet, in many studies of lag time to treatment of cancer patients, there is limited or no discussion about estimands that could be interpreted causally, and therefore that could be used in comparative assessments of treatment initiation policies or clinical guidelines. Here, we follow principles of the target trial emulation framework (11-13) to discuss a causally interpretable and clinically relevant estimand and a target trial protocol that could be used to quantify effects of lag time from diagnosis to oncologic treatment. In the next section, we describe the notation used throughout. Then, we define estimands that, we argue, are relevant in this context. Next, we propose the protocol for a target trial that would derive one of these estimands, and present simulations that illustrate its use. Although, for clarity, we focus on one type of lag time (from diagnosis to treatment), the approach described here could, with some adaptations, be used in the study of other lag times postulated to influence cancer outcomes.

## 2. Methods

### 2.1 Notation

We consider a study that includes patients diagnosed with a particular type of cancer. Participants are followed for a maximum time of *T* + 1 months; here, we use discrete time (e.g. months 0,1, …, *T*) to refer to the follow-up and timing of events. Further, we denote the binary treatment status at time *t* with *A*_*t*_ (1 = patient treated at time *t*; 0 = patient not treated at time *t*). We denote death status at month *t* with *Y*_*t*_ (1 = patient dead at time *t*; 0 = patient alive at time *t*). If *A*_*t*_ = 1 (*Y*_*t*_ = 1), then *A*_*t*_′ = 1 (*Y*_*t*_′ = 1) for *t* ≤ *t*^′^ ≤ *T*. Moreover, 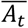 corresponds to treatment history relative to time *t*, that is, 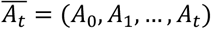 and 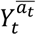 represents the potential outcome of *Y* at time *t* under a treatment strategy 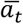.

## 3. Results

### 3.1 Causal estimands for the effect of lag time

Different causal questions, and hence estimands, can be considered in the context of studies on lag time to oncologic treatment. For instance, a causal estimand that could be targeted is the following:

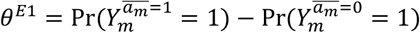

which contrasts counterfactual quantities in scenarios where participants are treated from time 0 versus not treated for the entirety of the follow-up *m* (*m* ≤ *T*) used to define the outcome; these two treatment strategies correspond to deterministic static strategies.(14) Although *θ*^*E*1^ is a valid causal estimand, for a value of *m* before which a substantial proportion of outcome events occurs, *θ*^*E*1^ does not capture the impact of treatment delay, but rather that of treatment versus non-treatment.

In fact, the term “lag time” implies a comparison of treatment strategies where initiation of treatment in one strategy occurs earlier than that under another strategy; this also suggests an informative causal question from a policy standpoint: if public health professionals had to decide on resource allocation, so that waiting times in cancer care could be reduced, they would be interested in quantifying the, potentially negative, effect that a treatment delay of *d* months might have on clinical outcomes. In this case, the following estimand might be of interest:

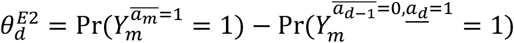

where 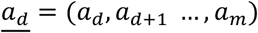 The estimand 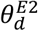 thus depends on the delay *d* that is being considered, and is a quantitative measure of the effect of delaying treatment by *d* months. We argue that 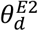 might be more relevant for the study of lag times than *θ*^*E*1^ and than the following estimand

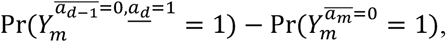

which quantifies the effect of initiating treatment at time *d* versus not initiating treatment; we note that the latter estimand is analogous to one of the estimands in Appendix S1.1 of (15), described in the different context of vaccine effectiveness.

We might also consider treatment strategies that use grace periods (16). For example, we could consider a treatment strategy that uses a grace period of 3 months: start treatment within 3 months after diagnosis. For simplicity, in the next section, we will not consider grace periods.

### 3.2 Target trial comparing strategies defined by timing of treatment initiation

The causal contrasts in the previous section could be estimated in a hypothetical trial that randomizes time of initiation of therapy. Although such trials might, in general, not be feasible, to clearly frame the causal questions that might be asked in observational studies on lag times, we consider the design of these trials. Below, we focus on the causal question

> “What is the effect, expressed in terms of survival at time *t*, of delaying treatment initiation by *d* months for the population of patients with diagnosis of a particular type of cancer and that require a particular type of therapy?”

Hence, our focus is on the causal estimand 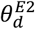. Specifically, we (i) discuss different components of the protocol of the relevant target trial, and (ii) outline how observational data might be used to emulate this trial – the two steps of the target trial emulation framework (17).

First, regarding the eligibility criteria for the target trial, eligible individuals would be those diagnosed with a particular cancer type during the recruitment window. If the causal question of interest refers to a particular therapeutic modality, investigators would need to restrict enrolment to patients who require curative therapy of a similar type (e.g. surgery or chemotherapy); one reason for studying lag time effects in respect to a particular treatment modality is the possibility that different treatment types might involve intrinsically different lag times. We note, however, that for some types of cancer and settings, this might be difficult, as it is possible that for some patients the delay becomes so critical that the disease may require more aggressive treatment combinations than it did at the time of diagnosis, due to a stage shift to a more advanced condition. In other words, it is conceivable that, in some contexts, treatment delay affects treatment modality itself. Similar eligibility criteria would be used in the emulation of the target trial; for that, date of cancer diagnosis would need to be available in the study dataset (e.g. from codes in medical registry databases). Note that although we chose to focus on the effect of treatment delays for the entire population of patients diagnosed with a particular cancer type, and possibly requiring a particular treatment modality, regardless of disease severity at diagnosis, it could be argued that the population that should be enrolled in such a target trial ought to include only those for whom it is realistically (in ethical terms) possible to delay treatment. On this view, we might want to restrict the study population to patients with, for instance, intermediate cancer staging and grading, and the causal estimand here would then be 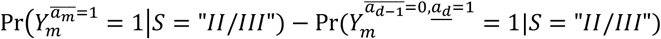, where *S* is a categorical variable indicating cancer staging, and "*II*/*III*" corresponds to Stages II or III.

As for the treatment strategies, we consider initiation of treatment during the first month after diagnosis (*t* = 0) versus treatment initiation at time *d*, with each value *d* corresponding to a different treatment strategy. Although in a clinical trial, the treatment strategy assigned to a participant is unambiguously defined from time 0, in an observational emulation of the trial, the treatment strategy defined by a lag time *d* would remain unrevealed until treatment is actually initiated at time *d*. For instance, if we consider treatment strategies defined by 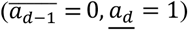, then when emulating the target trial with observational data, individuals who are not treated during the first month after diagnosis would have data compatible, at time 0, with multiple strategies where *d* > 0. For these participants, clones would be created in the dataset that would be assigned to different strategies compatible with data at time 0. Whenever a clone deviates from the assigned strategy, it would be artificially censored (18). For example, if, for a patient, treatment is initiated at time *t*, then clones of this patient under strategies with *d* > *t* would be censored at *t*. In **Figure 1**, we illustrate cloning of data from study participants and censoring of clones that deviate from treatment strategies; note, for instance, that for the individual who remained untreated until the time of death at *t* = 4, the death event would count for all treatment strategies with *d* > 4.

**Figure 1.**
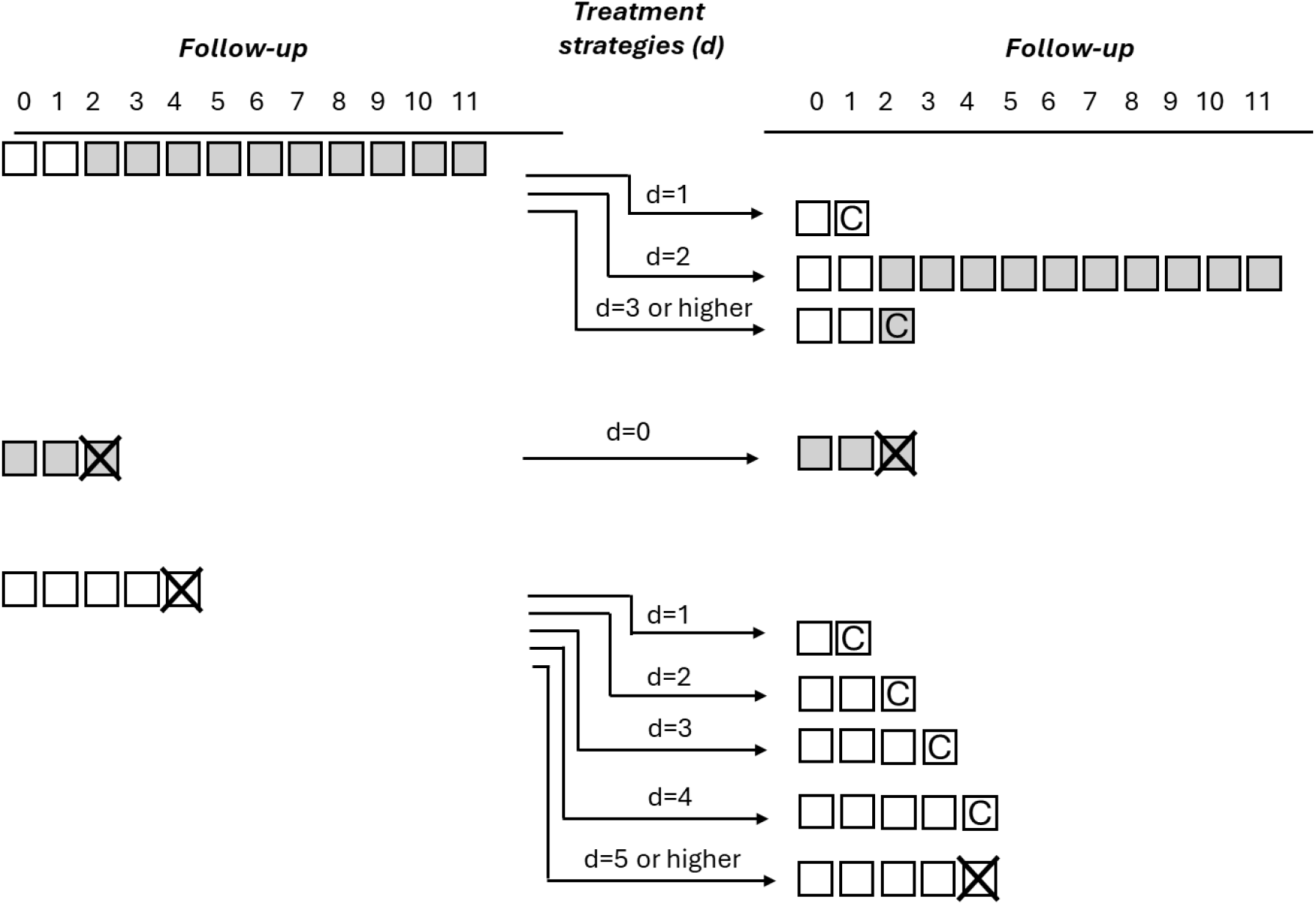
Illustration of the follow-up in an observational study and of clone-specific censoring. Hypothetical data from three study participants are shown in the left; each square represents a month of the follow-up. Treatment is denoted by grey, and death by an “X”. In the right side of the figure, we present information on clones for each participant; “C” indicates censoring.

A component of the target trial protocol that is definitionally different in the observational emulation is the assignment of treatment strategies. While in the target trial, assignment of treatment strategies would involve randomization, in the emulation, we need to assume no unmeasured confounding conditional on covariates, which here would imply sequential conditional exchangeability for censoring, allowing to address selection bias from artificial censoring of clones. Note that in our context both the ideal trial (19) and the target trial would be unblinded, as effects of treatment delay mediated through knowledge of treatment timing would be of intrinsic interest.

Moreover, for both the target trial and its observational emulation, time zero of follow-up would coincide with cancer diagnosis, which, in the target trial, would also coincide with treatment strategy assignment; and follow-up would end at time *m* for all treatment strategies or at the time of death.

Finally, the statistical analysis of the trial would involve a comparison of the outcome *Y*_*m*_ in different treatment arms; both intention-to-treat and per protocol analyses could be performed in the target trial. In the observational emulation, we would need to focus on the per protocol causal contrast, and would account for the potential selection bias introduced by censoring (**Figure 1**) using inverse probability weighting (18, 20, 21) (see Appendix 2 in (22) for a discussion on relevant assumptions).

### 3.3 Numerical example

Here, we illustrate the approach described above with an application. We simulated a hypothetical observational study in which patients were followed from diagnosis of an aggressive form of cancer until the end of follow-up, six months later, or until death, whichever occurred earlier. Our objective was to estimate the effect of starting treatment on month 0 versus month 2 after diagnosis; the outcome of interest was death in the first six months of follow-up. The simulation approach, with the assumed parameter values, is described in the *Appendix* (*Simulation approach for the numerical example* section). Briefly, the underlying data generating mechanism involved a fixed probability of treatment initiation at each month, and the hazard of death was assumed to be reduced by treatment.

In **Table 1**, data from two hypothetical study populations are presented: one in which the Waiting Time Paradox is absent, and another where this bias is present and individuals with more severe disease have a higher probability of treatment initiation. For Scenario I of **Table 1**, a lower proportion of participants treated on month 0 are alive 6 months after diagnosis compared to those who initiate treatment at month 2 (0.84 versus 0.88, respectively; difference -0.04, 95% confidence interval [CI]: -0.05, -0.03). This comparison is affected by immortal time bias (23), as participants starting treatment at month 2 could not, by definition, have developed the outcome during the first two months. A similar pattern is observed in Scenario II, where the proportions of participants who initiated treatment at month 0 versus month 2 and who were alive at month 6 were: 0.57 versus 0.69 for patients with more severe disease, and 0.84 versus 0.89 for patients with less severe disease; in the total population, these proportions were 0.66 versus 0.79 (difference -0.14, 95% CI: -0.15, -0.13). Note that in these comparisons, data from participants who did not receive treatment or who initiated treatment at month 1 or after month 2 are not included. Now, if we emulate the target trial described in the previous section using these data, then each participant might contribute to multiple treatment strategies defined based on lag time to treatment; such analyses require data cloning, censoring and weighting (18). For Scenario I, we estimated, using the inverse-probability-weighted Kaplan Meier method, that survival at 6 months was 0.84 versus 0.79 for treatment strategies with *d* = 0 versus *d* = 2 (difference 0.05, 95% CI: 0.04, 0.06); this coincides with the true values based on a simulation in which the entire population received these strategies. For Scenario II, survival at 6 months was 0.70 versus 0.63 for treatment strategies *d* = 0 versus *d* = 2 (which, again, coincides with the true values); the difference in survival was 0.07, 95% CI: 0.06, 0.08.

**Table 1.**
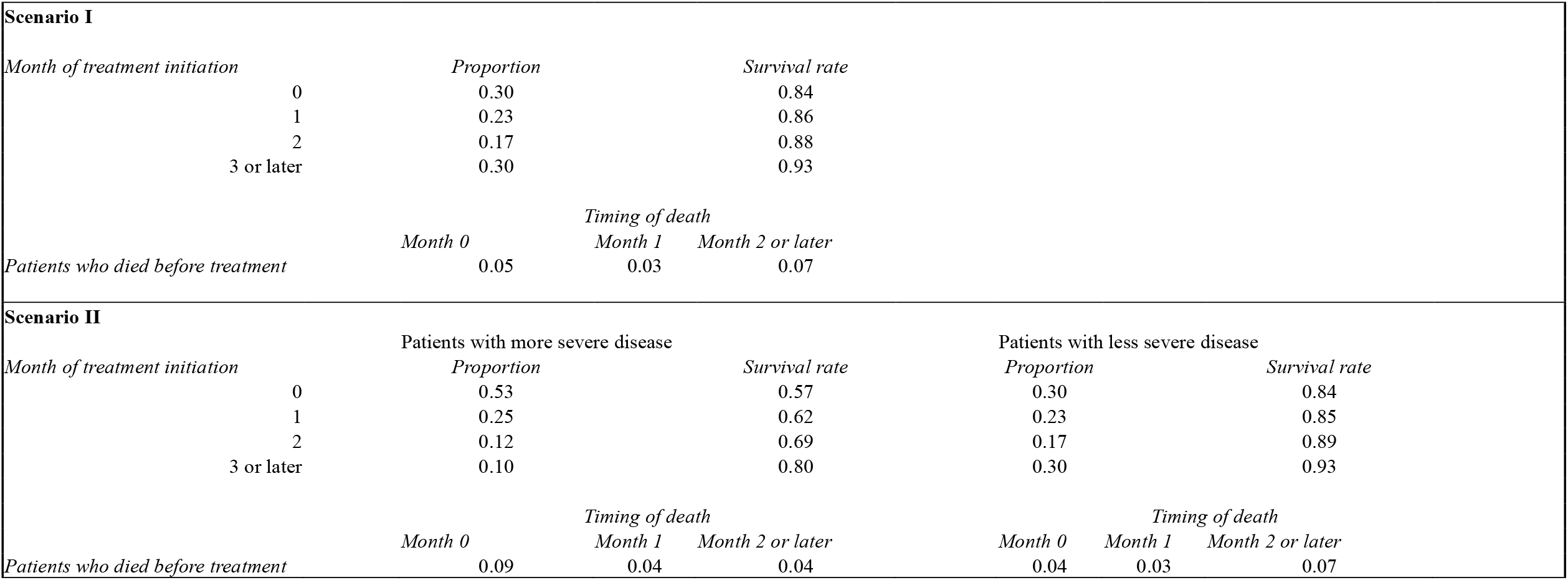
Numerical example. For Scenario I, we present data for a population without indication bias; data are stratified by month of treatment initiation (proportions refer to the group that received treatment, and not to the total population). The column “Survival rate” represents the proportions of patients alive at month 6. Some patients died before treatment initiation; the month of death is indicated for these patients (here, proportions refer to the total population). In Scenario II, we assume disease severity influences both timing of treatment initiation and the clinical outcome; in this scenario, proportions are presented for each severity category. For each scenario, we simulated data for 50,000 participants, which allows precise estimates. For example, for Scenario I, the difference between survival at 6 months for treatment strategies with *d* = 0 versus *d* = 2 is 0.05 (95% CI 0.04, 0.06; confidence interval was calculated from bootstrap samples), and for Scenario II, this difference is 0.07 (95% CI 0.06, 0.08). In reality, sample sizes might be considerably smaller, which could also lead to positivity issues as few patients begin treatment at any given time; this could be alleviated by using grace periods or coarsening intervals.

## 4. Discussion

Use of the target trial emulation framework has led to significant improvements in epidemiologic practice aimed at addressing causal questions, and multiple observational studies in cancer epidemiology have employed this framework (24-27). In this paper, we leverage the clarity provided by the target trial emulation framework to propose an estimand and a hypothetical trial design that might be useful when the scientific question of interest relates to the negative impact of treatment delays on oncologic outcomes. Indeed, while there have been many studies on lag times, not only between diagnosis and treatment, but also between other steps in cancer care (see review by Tope and colleagues (6)), we argue that the approach described here, and potentially other approaches based on this framework, would allow investigators to estimate quantities that are easier to interpret and more directly relevant to policy making. In fact, we note that a similar approach was used by Garcia-Albeniz and colleagues (28) to study the optimal timing of androgen deprivation therapy in prostate cancer patients with relapse, although in that study, instead of an alternative strategy defined in terms of a particular delay, the authors considered a “deferred” treatment strategy that was defined based on clinical criteria.

As indicated above, the type of question considered here is, in many respects, distinct from comparisons of treatments for which timing of initiation is assumed to approximately coincide. One of the differences relates to the fact that, in the observational emulation of target trials, strategies defined based on lag time to treatment might not be distinguishable at time 0, whereas in comparisons of different therapeutic approaches (e.g. different chemotherapy regimens), they likely are. Another potential complication in studies of the influence of lag time is that longer delays to therapy mean that treatment might start at later stages of the natural disease progression. An analogous problem was studied for the initiation of antiretroviral therapy in patients with HIV infection (29); in that study, Cole and colleagues argued that while a trial would capture the events for fast progressors in the arm that corresponds to deferred treatment, an observational study with start of follow-up defined by treatment initiation would not account for these factors. As discussed in the study by Cain and colleagues (18), where an approach similar to the one proposed here was used, the target trial formulation of treatment strategies avoids this problem.

In addition to allowing precise articulation of causal questions, emulations of target trials also avoid design-related biases. This approach, however, might still be affected by confounding, and hence, above, we assumed that a database would be available with information on eligibility criteria and confounders (including possible time-varying confounders). Alternatively, prospective studies could be designed to collect information on a sufficient set of variables that would allow for the control of confounding. Relatedly, while we considered disease severity at the time of diagnosis as a possible source of bias, other factors might be important, including age and presence of comorbidities. In real-world studies, the statistical method used in the analysis (e.g. inverse probability weighting) would thus need to account for these factors to enable a quasi-experimental framework to infer causality.

Finally, it is important to clarify that although the comparisons above were defined relative to *d* = 0, that is, to the treatment strategy 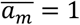, it might be the case that treatment initiation in the month following diagnosis is not feasible. As mentioned in a previous section, treatment strategies with grace periods (for instance, the strategy of starting treatment any time during the first 3 months after diagnosis) could be useful in this context. Alternatively, comparisons could be defined in reference to a delay *d* that would be minimal among those that are logistically feasible in a particular setting.

In summary, estimating the effects of lag time to treatment of cancer patients would help policymakers to weigh the benefits of interventions that would minimize such delays. Meaningful analyses of these effects require first identifying a relevant and precise causal question and a corresponding estimand, and then describing the protocol of a target trial that would answer this question. By doing just this, we hope this paper will facilitate the design of causal analyses on lag times and improve oncologic outcomes. Moreover, some of the considerations presented here could be used as a starting point by investigators to ensure that their studies on lag time comply with the recently published Transparent Reporting of Observational Studies Emulating a Target Trial (TARGET) Statement (17).

## Data Availability

All data produced in the present work are contained in the manuscript.

## Appendix

### Table of contents

Simulation approach for the numerical example

Additional information on simulations presented in **Table 1**

References

#### Simulation approach for the numerical example

To generate the data in **Table 1**, we first defined the probability of death by the end of the follow-up in the absence of treatment, 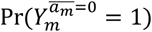 in simulations presented in the main text, we assume this probability to be 0.30. For simplicity, we assume that the hazard of death was stable during the follow-up:

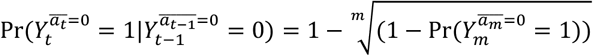

We modelled treatment initiation with:

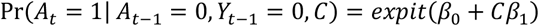

where *A*_*t*_ is a binary variable indicating treatment at time *t*; *β*_0_ = *logit*(0.20) and for Scenario II (**Table 1**) *β*_1_ = log (3) (for Scenario I, *β*_1_ = log(1)); *C* is a binary variable indicating disease severity, and in Scenario II, the probability of presenting more severe disease is assumed to be 0.50.

The impact of disease severity and treatment on the hazard of death was modelled as:

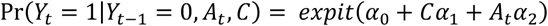

where *α* = *logit*(0.06) (based on 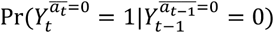 defined above), and *α*_1_ and *α*_2_ are parameters representing log-odds ratios. We assumed *α*_2_ = log (0.50). In Scenario I,*α*_1_ = log (1), and in Scenario II, *α*_1_ = 1.14 was calculated so that 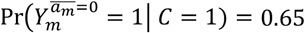.

Given the above, the true values for the quantities determining the causal contrasts of interest are:

Scenario I

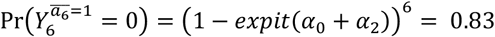

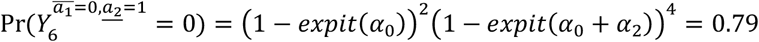

Scenario II

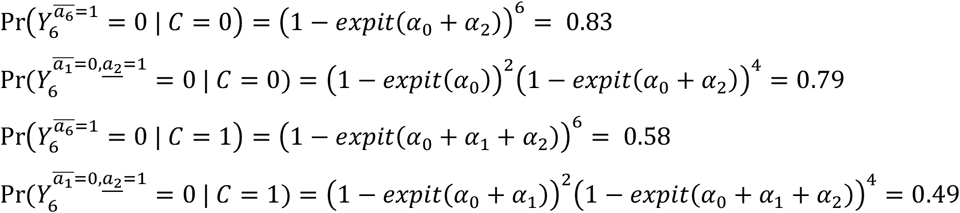

For analyses that emulate a target trial, data from participants were cloned for each treatment strategy with which they were consistent during follow-up; clones were then censored whenever data deviated from the corresponding strategy. To account for selection bias introduced by censoring, we used stabilised inverse probability of censoring weights (1), where terms were estimated using logistic regressions applied to the original dataset, without clones. Weights were used in Kaplan-Meier analyses to estimate survival curves.

#### Additional information on simulations presented in Table 1

For Scenario I of **Table 1**, 66.5% of participants received treatment during the six months following diagnosis, and 23.2% died during follow-up. Of the 33.5% that were not treated in the first six months, nearly half (15.2%) died during the follow-up. In the expanded dataset with data clones, 10,050/50,000 individuals were assigned to the treatment strategy corresponding to *d = 0*, and 39,950/50,000 individuals, to treatment strategies *d* = 1 and *d* = 2 (we did not consider strategies with *d* > 2).

In Scenario II, 74.0% of participants received treatment during the six months following diagnosis, and 35.9% died during follow-up. In the expanded dataset with clones, 15,762/50,000 individuals were assigned to the treatment strategy corresponding to *d = 0*, and 34,238/50,000 individuals, to treatment strategies *d* = 1 and *d* = 2.

## References

1. Tope PR, Goncalves BP, El-Zein M, Franco EL. The health related impact of disruptions in cancer care and the Waiting Time Paradox. Am J Epidemiol. 2025.

2. Gorey KM, Luginaah IN, Holowaty EJ, Fung KY, Hamm C. Wait times for surgical and adjuvant radiation treatment of breast cancer in Canada and the United States: greater socioeconomic inequity in America. Clin Invest Med. 2009;32(3):E239–49.

3. Molinié F, Leux C, Delafosse P, Ayrault-Piault S, Arveux P, Woronoff AS, et al. Waiting time disparities in breast cancer diagnosis and treatment: A population-based study in France. The Breast. 2013;22(5):810–6.

4. Crawford SC, Davis JA, Siddiqui NA, de Caestecker L, Gillis CR, Hole D, et al. The waiting time paradox: population based retrospective study of treatment delay and survival of women with endometrial cancer in Scotland. Bmj. 2002;325(7357):196.

5. Dang-Tan T, Trottier H, Mery LS, Morrison HI, Barr RD, Greenberg ML, et al. Determinants of delays in treatment initiation in children and adolescents diagnosed with leukemia or lymphoma in Canada. Int J Cancer. 2010;126(8):1936–43.

6. Tope P, Farah E, Ali R, El-Zein M, Miller WH, Franco EL. The impact of lag time to cancer diagnosis and treatment on clinical outcomes prior to the COVID-19 pandemic: A scoping review of systematic reviews and meta-analyses. eLife. 2023;12:e81354.

7. Mullen CJR, Barr RD, Franco EL. Timeliness of diagnosis and treatment: the challenge of childhood cancers. Br J Cancer. 2021;125(12):1612–20.

8. Allgar VL, Neal RD. Delays in the diagnosis of six cancers: analysis of data from the National Survey of NHS Patients: Cancer. Br J Cancer. 2005;92(11):1959–70.

9. Neal RD. Do diagnostic delays in cancer matter? Br J Cancer. 2009;101 Suppl 2(Suppl 2):S9–s12.

10. Dang-Tan T, Franco EL. Diagnosis delays in childhood cancer: a review. Cancer. 2007;110(4):703–13.

11. Hernan MA, Wang W, Leaf DE. Target Trial Emulation: A Framework for Causal Inference From Observational Data. JAMA. 2022;328(24):2446–7.

12. Hernan MA, Dahabreh IJ, Dickerman BA, Swanson SA. The Target Trial Framework for Causal Inference From Observational Data: Why and When Is It Helpful? Ann Intern Med. 2025;178(3):402–7.

13. Hernan MA, Robins JM. Using Big Data to Emulate a Target Trial When a Randomized Trial Is Not Available. Am J Epidemiol. 2016;183(8):758–64.

14. Hernán MA, Robins JM. Causal Inference: What If: Boca Raton: Chapman & Hall/CRC; 2020.

15. Boyer C, Lipsitch M. Emulating target trials of postexposure vaccines using observational data. Am J Epidemiol. 2024.

16. Wanis KN, Sarvet AL, Wen L, Block JP, Rifas-Shiman SL, Robins JM, et al. Grace periods in comparative effectiveness studies of sustained treatments. J R Stat Soc Ser A Stat Soc. 2024;187(3):796–810.

17. Cashin AG, Hansford HJ, Hernan MA, Swanson SA, Lee H, Jones MD, et al. Transparent Reporting of Observational Studies Emulating a Target Trial-The TARGET Statement. JAMA. 2025.

18. Cain LE, Robins JM, Lanoy E, Logan R, Costagliola D, Hernan MA. When to start treatment? A systematic approach to the comparison of dynamic regimes using observational data. Int J Biostat. 2010;6(2):Article 18.

19. Moreno-Betancur M, Wijesuriya R, Carlin JB. The Ideal Trial: Defining Causal Estimands that Balance Relevance and Feasibility in Target Trial Emulations and Actual Randomized Trials. Epidemiology. 2026;37(2):153–62.

20. Rudolph JE, Schisterman EF, Naimi AI. A Simulation Study Comparing the Performance of Time-Varying Inverse Probability Weighting and G-Computation in Survival Analysis. Am J Epidemiol. 2023;192(1):102–10.

21. Hernán MA. How to estimate the effect of treatment duration on survival outcomes using observational data. BMJ. 2018;360:k182.

22. Cole SR, Hernán MA. Constructing inverse probability weights for marginal structural models Am J Epidemiol. 2008;168(6):656–64.

23. Hernan MA, Sauer BC, Hernandez-Diaz S, Platt R, Shrier I. Specifying a target trial prevents immortal time bias and other self-inflicted injuries in observational analyses. J Clin Epidemiol. 2016;79:70–5.

24. McGee EE, Hernan MA, Giovannucci E, Mucci LA, Chiu YH, Eliassen AH, et al. Estimating the Effects of Lifestyle Interventions on Mortality Among Cancer Survivors: A Methodologic Framework. Epidemiology. 2025;36(5):705–18.

25. Garcia-Albeniz X, Hsu J, Bretthauer M, Hernan MA. Effectiveness of Screening Colonoscopy to Prevent Colorectal Cancer Among Medicare Beneficiaries Aged 70 to 79 Years: A Prospective Observational Study. Ann Intern Med. 2017;166(1):18–26.

26. Garcia-Albeniz X, Hsu J, Hernan MA. The value of explicitly emulating a target trial when using real world evidence: an application to colorectal cancer screening. Eur J Epidemiol. 2017;32(6):495–500.

27. Chesang C, Sharples LD, Gray CM, Nossiter J, van der Meulen J, Cowling TE, et al. Emulating an existing trial of treatments for prostate cancer using real-world data: challenges and lessons learned. J Clin Epidemiol. 2025;182:111767.

28. Garcia-Albeniz X, Chan JM, Paciorek A, Logan RW, Kenfield SA, Cooperberg MR, et al. Immediate versus deferred initiation of androgen deprivation therapy in prostate cancer patients with PSA-only relapse. An observational follow-up study. Eur J Cancer. 2015;51(7):817–24.

29. Cole SR, Li R, Anastos K, Detels R, Young M, Chmiel JS, et al. Accounting for leadtime in cohort studies: evaluating when to initiate HIV therapies. Stat Med. 2004;23(21):3351–63.

## References

1. Cain LE, Robins JM, Lanoy E, Logan R, Costagliola D, Hernan MA. When to start treatment? A systematic approach to the comparison of dynamic regimes using observational data. Int J Biostat. 2010;6(2):Article 18.

